# How group structure impacts the numbers at risk for coronary artery disease: polygenic risk scores and non-genetic risk factors in the UK Biobank cohort

**DOI:** 10.1101/2023.07.28.23292953

**Authors:** Jinbo Zhao, Adrian O’Hagan, Michael Salter-Townshend

**Affiliations:** Insight Centre for Data Analytics, University College Dublin, Belfield, Dublin, D04V1W8, Ireland; School of Mathematics and Statistics, University College Dublin, Belfield, Dublin, D04V1W8, Ireland

**Keywords:** UK Biobank, Coronary artery disease, Polygenic risk score, Pooled cohort equation risk, Group structure

## Abstract

The UK Biobank is a large cohort study that recruited over 500,000 British participants aged 40-69 in 2006-2010 at 22 assessment centres from across the UK. Self-reported health outcomes and hospital admission data are two types of records that include participants’ disease status. Coronary artery disease (CAD) is the most common cause of death in the UK Biobank cohort. After distinguishing between prevalence and incidence CAD events for all UK Biobank participants, we identified geographical variations in age-standardised rates of CAD between assessment centres. Significant distributional differences were found between the pooled cohort equation scores of UK Biobank participants from England and Scotland using the Mann-Whitney test. Polygenic risk scores of UK Biobank participants from England and Scotland and from different assessment centres differed significantly using permutation tests. Our aim was to discriminate between assessment centres with different disease rates by collecting data on disease-related risk factors. However, relying solely on individual-level predictions and averaging them to obtain group-level predictions proved ineffective, particularly due to the presence of correlated covariates resulting from participation bias. By using the Mundlak model, which estimates a random effects regression by including the group means of the independent variables in the model, we effectively addressed these issues. In addition, we designed a simulation experiment to demonstrate the functionality of the Mundlak model. Our findings have applications in public health funding and strategy, as our approach can be used to predict case rates in the future, as both population structure and lifestyle changes are uncertain.

## Introduction

### Coronary artery disease

Coronary artery disease (CAD), sometimes referred to as coronary heart disease (CHD) or ischemic heart disease, is a common heart condition that occurs when the blood and oxygen supply to the heart muscle is inadequate, and is one of the leading causes of morbidity and mortality in the United Kingdom, the United States and worldwide (e.g., Cheema *et al*. (2022), Shahjehan and Bhutta (2022)). Many environmental factors including smoking, unhealthy diet, alcohol intake, obesity, hypertension, diabetes mellitus, and lack of physical activity, have impact on the development of CAD (Mack and Gopal 2016). Family history of cardiovascular disease has been extensively researched as a standalone risk factor for CAD both in the short and long term (e.g., Lloyd-Jones *et al*. (2004), Bachmann *et al*. (2012)).

Several risk scores have been proposed to estimate the future cardiovascular risk (e.g., over the next 10 years) for currently healthy people, such as the Framingham risk score (FRS) (D’Agostino Sr et al. 2008), QRISK3 (risk score using the QRE-SEARCH database) (Hippisley-Cox et al. 2017) and pooled cohort equation (PCE) scores (Goff et al. 2014). These scores combine the effects of multiple carefully selected non-genetic risk factors into a single score, and the effect of each risk factor or interaction term is estimated through sophisticated statistical analysis. Family history is included in QRISK3, but not in the other two scores. Those overall risk scores are clinically meaningful. For example, if a currently healthy person is diagnosed with a PCE-estimated 10-year cardiovascular disease risk exceeding 7·5%, they will be advised to take statin therapy to reduce their future cardiovascular risk after consultation with their doctor in the US (Vasan and Van den Heuvel 2022).

Our understanding of the genetic structure of CAD is also increasing with the development of gene sequencing and analysis technologies. Genotyping microarrays designed to capture most common inter-individual genetic variation provide the basis for genome-wide association studies (GWAS) (Khera and Kathiresan 2017). Since the first GWAS on CAD reported three common variants associated with increased risk of CAD, more than 200 causal variants have been identified in association with the development of CAD (Aragam *et al*. 2022). Apart from the association signal between the causal variants and the phenotypes, causal variants also have biological effects on the phenotypes (Hormozdiari *et al*. 2015). GWASs also detect many genetic variants that have no biological effect but are statistically significant for phenotypes (Visscher *et al*. 2017).

For many years, the field of genetics has focused extensively on efforts to predict human diseases and traits, and polygenic risk scores (PRS) have the potential to be useful in clinical settings, particularly in the context of specific purposes and conditions (Ogbunugafor and Edge 2022). PRS is a tool that translates personal genetic information into real numbers that can be interpreted as an individual’s genetic risk for a particular disease. There is already compelling evidence indicating its effectiveness in predicting the risk of CAD. For example, the utility of CAD-PRS as an independent risk factor for predicting the risk of CAD has been widely recognised and discussed (e.g., Dikilitas *et al*. (2022)).

The genetic risk score for CAD can be represented by polygenic risk scores (PRS), which combines the effects from both causal and significant variants. PRS is a tool that translates personal genetic information into real numbers that can be interpreted as the individual-level genetic risk of a specific disease. The utility of CAD-PRS as an independent risk factor to predict the risk of CAD has been widely identified and discussed (e.g., Dikilitas *et al*. (2022)).

The combination of genetic and non-genetic risk factors increases the predictive power at the individual level. Elliott *et al*. (2020) calculated CAD-PRS and PCE scores for their study participants and compared the predictive power of risk factors alone and combined. They found that the overestimation of risk by PCE scores could be corrected by adding CAD-PRS to the model. Comparing the model with only PCE to the model with PCE and PRS, when using a risk threshold of 7.5%, the latter improved net reclassification 4.4% for cases and -0.4% for controls. Incorporating family history and PRS can improve the accuracy of predicting CAD risk in both real-world and simulation study settings (e.g., Hujoel *et al*. (2022) Zhao *et al*. (2023)).

### Geographical variations in cardiovascular disease prevalence across the UK

Cardiovascular disease (CVD) is the term for all types of diseases that affect the heart or blood vessels and CAD is the most common type of CVD. Within the UK, the higher prevalence of cardiovascular disease (CVD) in Scotland than in England has been repeatably observed (e.g., Lawlor *et al*. (2003), Bhatnagar *et al*. (2016)). The recent epidemiology study conducted by Cheema *et al*. (2022) shows the age standardised CVD mortality rate differences in 2019 across 13 UK regions/nations, including the East Midlands, East England, London, Yorkshire and the Humber, Wales and Scotland. Among those regions, Scotland has the highest mortality rate per 100,000 for CVD for all ages.

Environmental and genetic risk factors can both contribute to geographical variations in CVD (e.g., Lawlor *et al*. (2003), Peasey *et al*. (2006), Ding and Kullo (2009)). For example, Lawlor *et al*. (2003) concluded that age distribution, socioeconomic status, and health service utilization were the main causes of geographical variation, as well as differences in risk factors associated with CVD, including smoking, hypertension status, blood pressure and cholesterol levels. Ethnic-specific differences in the genetic architecture of CAD have been widely proposed and explored, and different novel disease-susceptibility loci have been identified in different populations (Miyazawa and Ito 2021).

Geographical variations in CAD prevalence were reflected in the UK Biobank (UKB) participants, with CAD prevalence of 7.73% in England UKB participants and 9.06% in Scotland UKB participants (Yang et al. 2021). Yang *et al*. (2021) conducted a study on UKB participants to explore whether environmental or genetic factors could explain the regional CAD prevalence differences. They calculated the FRS, QRISK3 and PRS for CAD risk and concluded that neither FRS, QRISK3 or PRS could explain the higher CAD prevalence in Scotland. They used Pearson’s Chi-squared test and the two-tailed Mann-Whitney test for statistical analysis. However, because they observed significant differences in the distribution of individual risk alleles, they concluded that the genetic architecture of a common disease could be different for geographically and ethnically closely related populations.

### Study aim

Genetic and non-genetic risk factors working together can improve the prediction of CAD risk at the individual level (e.g., Elliott *et al*. (2020), Hujoel *et al*. (2022)), but few studies have used them jointly to estimate the number of risks at the regional/country level. In this study, we are interested in comparing, analyzing and predicting the risk of CAD at the regional/country level employing the UKB participants. Study participants and regional selection are explained in Section Study participants and CAD events, followed by the test methods used to compare the distribution of PCE and a different set of CAD-PRS at the group level. Section Results for UKB assessment centres contains predictions of the number of people at risk for CAD at the regional level using a generalized linear model regressed on PCE and PRS, with a poor ability to distinguish between high and low case rate groups (Section Results for UKB assessment centres). The results in Ascertainment bias confounds group rate estimation show that it is ascertainment bias that confounds group rate estimation. Participation bias is common in population-based cohort studies, including the UKB study, and can bias the results of genetic epidemiology studies (Schoeler *et al*. 2023). The Mundlak model (Dieleman and Templin 2014) is used to eliminate bias and improve efficiency. The updated results show that the Mundlak model is valid. A simulation experiment (Section How the Mundlak model works and Section Simulation results) is designed to explain how the Mundlak model works.

## Methods

### Study participants and CAD events

#### UKB resources and health outcomes records

The data set for our work was created using data fields provided by the UKB resources under Application Number 59528. The UKB study (Sudlow *et al*. 2015) recruited half million UK participants aged 40-69 from across the UK during 2006-2010 for the baseline assessments. Over 70% of all UKB participants are from England, less than 10% are from Scotland, and the rest are from Wales, Northern Ireland and other regions. The baseline assessments were conducted at 22 assessment centres in Scotland, England and Wales and consisted of a five-part assessment process lasting 2-3 hours. The process included written consent, answering touch screen questionnaires, face-to-face interviews with a study nurse, measurements like hand grip and bone density, and the sample collection of blood, urine and saliva. The collected samples were used for gene sequencing and biochemical markers measurement, with various types of genetic data released since May 2015 (Bycroft *et al*. 2018).

UKB resources provide two types of record containing participants’ disease status, self-reported health outcomes, and hospital inpatient data. Participants were asked to report their health outcomes during the baseline assessment, including the type of disease(s) and the date(s) of onset. Additionally, UKB also keeps track of each participant’s hospital inpatient data, including hospital admissions information and date of admission, diagnosis during admission, procedures and discharge information. For example, hospital inpatient data for UKB participants from England are provided by the Data Access Request Service (DARS), managed by National Health Service (NHS) digital, and provides hospital inpatient admissions data for English participants. Inpatient data for participants from Wales and Scotland are provided via different partnerships. The UKB resources have over ten thousand data fields, with more arriving all the time. Those data fields can be assigned to several categories, such as physical measurements, lifestyle, cognition and hearing, physical activity, imaging, biomarkers and genetics. Hospital diagnoses information accounts for almost half of all UKB data fields (Madakkatel et al. 2021). UKB participants’ health outcomes are accessed by different coding systems for self-reported records and for the hospital inpatient records. Detailed, self-reported health outcomes are recorded separately for cancer and non-cancer conditions using UKB designed data-coding. All clinical data in the hospital inpatient data are coded according to the World Health Organization’s International Classification of Diseases (ICD) and all operations and procedures in the hospital inpatient data are coded according to the Office of Population, Censuses and Surveys: Classification of Interventions and Procedures (OPSC) (UK Biobank: hospital inpatient data).

#### CAD definition

To identify UKB participants diagnosed with CAD, CAD codes within self-reported and hospital inpatient records need to be determined first. There is no precise definition of which diseases should be included in determining the onset of CAD for UKB participants. Our study followed the CAD definition from Elliott *et al*. (2020). In detail, six different categories were searched to determine CAD events, including ICD-10, ICD-9, OPCS-4, non-cancer illness code, operation code and the vascular/heart problems data field. The CAD definition is in Supplementary Table 1 and the related UKB data fields are in Supplementary Table 2 in the Supplementary Material.

We defined any CAD events that happened before the date of joining the UKB for the initial assessment as prevalence CAD, and any events that happened after joining the UKB as incidence CAD. Some participants had more than one CAD events in their records, either one category with at least two different types of CAD events, or more than 2 categories of CAD events. For those cases, we compared the dates for multiple events and kept the earliest CAD event in this study. ICD-10, ICD-9 and OPCS-4 have the CAD date, while the other three have the CAD onset age in integer values. There may be some bias in converting the date of CAD onset to age at CAD onset to determine the first CAD event, as the date of birth of the UKB participants was not available in this study, only the year of birth.

#### UKB assessment centres to represent geographical regions

After we identified the prevalence and incidence of CAD events, we calculated the age-standardized prevalence in 2010 and 2021 across the assessment centres. Of all 22 centres, only 2 were located in Scotland, the rest were in England and Wales, and one of the centres in England was a pilot centre for only the first month of the overall baseline assessment period and had a relatively small number of participants. The UKB team sent invitation letters to people who were predominantly located in urban areas and lived near any of the UKB assessment centres (Alten *et al*. 2022). Therefore, it is reasonable to use the assessment centre as a geographic location to compare CAD morbidity. To obtain the age-standardized prevalence rates, we also used the 2013 European Standard Population as in Cheema *et al*. (2022).

### Genetic and non-genetic risk scores

#### CAD-PRS set selection

The basis of PRS is that for most common diseases, their inheritance involves many common genetic variants with small effects, and combining those effects together has the ability to distinguish risk groups. The calculation process for PRS is complex and beyond the scope of this paper. Interested readers can learn more from Choi *et al*. (2020). The baseline function for PRS using additive genetic models summarizes the effects of a set of significant genetic variants, with the number of genetic variants varying from hundreds to several millions. Various PRS methods have been developed aimed at determining the set of variants included in the baseline calculation (e.g. Chang *et al*. (2015), Ge *et al*. (2019)) and/or to estimate the magnitude of the effect (e.g., Vilhjálmsson *et al*. (2015), Mak *et al*. (2017)).

Among various PRS methods available for inspection, we chose to compare the CAD-PRS set calculated via the method LDpred2 (Privé *et al*. 2020a) with the CAD-PRS set provided by the UKB (Thompson *et al*. 2022). LDpred2 infers the posterior mean effect size for each genetic variant by using a prior on effect sizes and linkage disequilibrium (LD) information from an external reference panel. LDpred2 can also estimate the proportion of significant genetic variants and heritability explained by selected variants. LDpred2 claimed that its method beat other common methods after testing on UKB participants and our reproduced results, as well as results from Aragam *et al*. (2022) both support their conclusion. Steps to calculate CAD-PRS using the LDpred2 method are in Appendix LDpreds CAD-PRS calculation. The UKB resources category 300 provides access to standard PRS and enhanced PRS for 28 diseases (including CAD) and 25 quantitative traits, with the standard set (centred and variance-standardised) calculated for all participants in the UKB using algorithms trained on external data only and the enhanced set calculated for a subgroup of 104,231 individuals in UKB trained on external data and a separate subgroup of UKB (Thompson *et al*. 2022). LDpred2 restricts its usage on samples with the same ancestry (White British), while Thompson *et al*. (2022) built their PRS algorithms using a Bayesian approach, combing data across multiple ancestries.

We compared the predictive performance of these two sets of CAD-PRS using AUC, the area under the receiver operator characteristics curve, which measures the discrimination concordance between risk scores and binary outcomes (Huang and Ling 2005). The AUC between each one of 2 sets of PRS and the 2 definitions of CAD phenotypes were calculated using *R* function *AUCBoot* from the *bigstatsr* package (Privé *et al*. 2018) and results are shown on Table 1. The UKB CAD-PRS has slighter higher AUC values and covers more UKB participants, so this study chose the UKB CAD-PRS for use in our analysis.

**Table 1.**
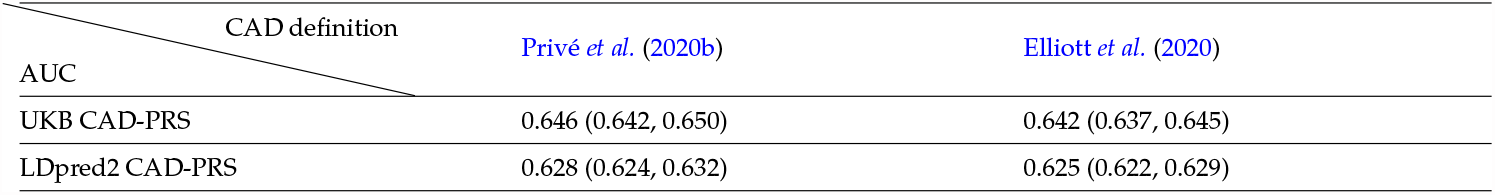
AUC comparison for two sets of CAD-PRS.

#### Calculation of pooled cohort equation scores

The American College of Cardiology (ACC) and the American Heart Association (AHA) developed pooled cohort equations (PCE) to estimate the composite endpoint of 10-year atherosclerotic cardiovascular (ASCVD) risk, with initial sex-specific and ethnicity-specific equations published in 2013 (Goff *et al*. 2014). Atherosclerosis is a common disease that occurs when a sticky substance called plaque builds up inside your arteries. ASCVD events include CAD, stroke, and peripheral artery disease (PAD) (DeFronzo and Ferrannini 1991). The PCE tool is a risk assessment method that has been developed based on data that can be easily collected by primary care providers and can be implemented in routine clinical practice. Carefully selected risk factors associated with CAD risk are included in PCE equations, including age, total and high-density lipoproteins (HDL) cholesterol levels, blood pressure, smoking status, diabetes mellitus and hypertension medication status. Log transformation and interaction terms are included in the equations. The PCE score is a single score that summarizes the effect using the parameters estimated by the proportional hazards model. The PCE scores for UKB participants have been studied widely, such as in Riveros-Mckay *et al*. (2021), Carter *et al*. (2022).

There are criteria for applying the PCE equation. Stone *et al*. (2014) points that it is not appropriate to estimate 10-year ASCVD using PCE scores for individuals with clinical ASCVD, or with LDL-C *≥*190 mg/dL, or people who are already in a statin benefit group.

We firstly identified UKB participants who already had an ASCVD event prior to joining the UKB, as the risk factors used to calculate PCE scores were collected at the baseline assessment visit. This study used the definitions of CVD from Elliott *et al*. (2020) to determine the prevalence CHD and stroke events, and the definition of PAD from Klarin *et al*. (2019), with relevant data fields from UKB, is in Supplementary Table 4. Following the CAD prevalence definition in Section Study participants and CAD events, the prevalence ASCVD events were determined by comparing their event onset dates with the date they joined the UKB. The corresponding events codes are in Supplementary Table 3. Of the 502,401 UKB participants, 35,308 were identified as participants with a first ASCVD epidemic event. An additional 155 participants did not have a corresponding date of first ASCVD epidemic event, but we still included them in the first ASCVD event group. In total, there are 35,887 UKB participants with first-ever ASCVD. Only 2 UKB participants had LDL-C ≥ 190 mg/dL during their initial assessment visit. Finally, we selected UKB participants who were already on statin therapy prior to joining UKB. We used the types of statin (atorvastatin, simvastatin, fluvastatin, pravastatin and rosuvastatin) listed by Carter *et al*. (2022).

This study employed the PCE coding provided in the supplementary material of Vasan and Van den Heuvel (2022) to calculate PCE risk scores. We also followed their additional criteria that PCEs were not applied for people with extreme total cholesterol(*>*320 or *<*130 mg/dL), high-density lipoprotein cholesterol (*>*100 or *<*20 mg/dL), or systolic blood pressure (*>*200 or *<*90 mm Hg). The risk factors associated with the UKB data fields are listed in Table S5 of File S1.

#### Study flow

The complete data set for this study included UKB participants who were eligible for PCE risk calculation and had CAD-PRS provided by UKB. In addition, body mass index (BMI) and Townsend deprivation index (TDI) were also extracted from the UKB resource for those participants, as BMI has been recognised as a risk factor that could aid the predictive power of PRS (e.g., Alten *et al*. (2022)) and TDI, a measure of social deprivation, has impact on the mortality of cardiovascular disease (e.g., Ford and Highfield (2016)). Figure 1 is the flow chart of obtaining this full data set for the following analysis. The complete data set had a total of 263,087 UKB participants, with 8,458 participants developing CAD after enrolling in the UKB and the remaining 254,629 participants remaining CAD-free.

**Figure 1.**
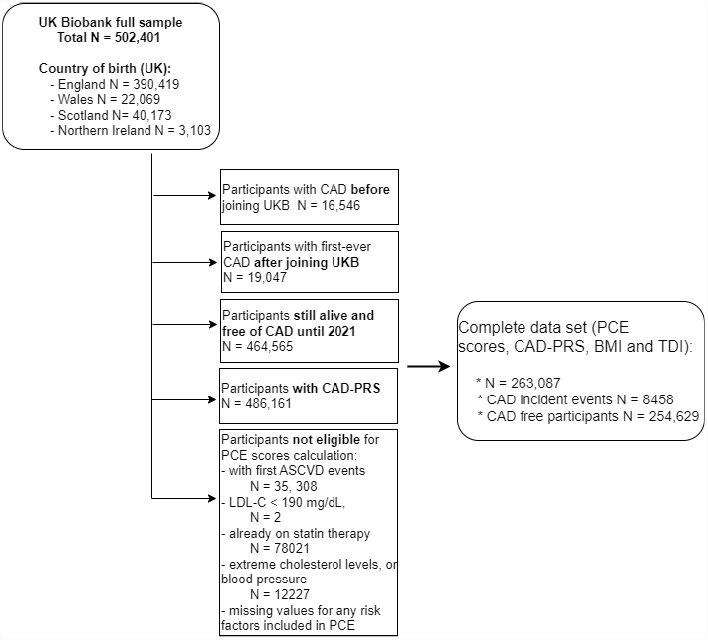
Study flow chart to generate the complete data set for further analysis. PCE denotes pooled cohort equation; PRS denotes polygenic risk score; BMI denotes body mass index; TDI denotes Townsend deprivation index; ASCVD denotes atherosclerotic cardiovascular; LDL-C denotes low-density lipoprotein cholesterol.

### Statistical tests

We examined the difference in PCE scores between England and Scotland, using a two-tailed Mann-Whitney test (Yang *et al*. 2021). The Mann-Whitney test is a nonparametric test and checks if two samples come from the same distribution by comparing the probability of *X* being greater than *Y* with the probability of *Y* being greater than *X* after randomly selecting values from sets *X* and *Y*. The Mann-Whitney test is considered to be a test of population medians and is accompanied by equally important differences in shape, but the Mann-Whitney test cannot discern differences between two groups with the same median, but can discern different variances or shapes, as this test analyzes only the ranks (Hart 2001).

To compare PRS distributions between any two assessment centres, we employed a permutation test. In this study we used the assessment centre to represent the geographical region, but we noted that the number of people going to an assessment centre close to their address was much lower than the number of people in that area. When we do not have access to the PRS of everyone in the region, but still want to compare the distributions of PRS, permutation tests are useful (Irizarry and Love 2016). People with PRS in the highest polygenic risk group have a higher chance of developing the disease than people with average PRS scores. For example, Lewis and Green (2021) examined the ability of PRS to predict risk for CAD using genotype and phenotype data from UKB participants, as the highest polygenic risk group had twice the hazard ratio of the intermediate risk group. Therefore, when comparing the PRS distributions of two populations, we are more interested in looking at the tails or spread of the PRS distribution than just comparing the means or variances. This is another reason why we chose to use the permutation test.

The permutation test is a resampling and nonparametric test that does not make any assumptions about the distribution; in fact, a full permutation test encompasses all possible permutations, hence it is a Monte Carlo permutation test. The permutation test requires four main steps:

1. determine and calculate the statistic of interest (e.g., mean, median or variance);
2. combine groups together, retaining all data but randomly shuffling the groups’ labels, and then calculate the new statistic value;
3. repeat step 2 many times and keep a record of the new statistic values;
4. the p-value is the proportion of statistics from the real group lower than the statistics from the reshuffled groups.

We also performed permutation tests on the predicted disease risk using the liability threshold model (LTM). The LTM assumes that there is a hidden continuous disease liability *L* that determines the binary disease outcome, where *L* follows a standard normal distribution, and the binary outcome *D* = 1 if *L* exceeds a fixed threshold *T* and 0 otherwise. The threshold is determined by the prevalence *K* of the disease in the population using the relationship *T* = Φ^*−*1^(1 *K*), where Φ is the cumulative distribution function of the normal distribution (So et al. 2011). The total liability *L* is assumed to be split into two components, the measurable genetic component and the combination of environmental and unknown risk factors, while PRS can be used to represent the measurable genetic component (Zhao *et al*. 2023). If we assume that the variance explained by PRS is *V*, then the LTM suggests that *Cov*(*L, PRS*) = *Var*(*PRS*) = *V* and 𝔼 (*L*|*PRS* = *prs*_*j*_) = *prs*_*j*_ and *Var*(*L*|*PRS* = *prs*_*j*_) = 1*− V*. Then, using standard regression theory, we can calculate the probability of being a case given the value of PRS as *Pr*(*L > T*|*PRS* = *prs*_*j*_) = *Pr*(*L− prs*_*j*_ *> T − prs*_*j*_) = 1 *−* Φ(*T prs*_*j*_, 0, 1 *− V*) (So et al. 2011).

In this study, we denoted the CAD incidence rates from the complete data as *P*, and calculated the predicted probability of being a case after standardising the CAD-PRS. The reason for this approach is that LTM provides a framework for predicting the risk of developing the disease solely based on PRS values within a specific group. Thus, two cohorts (whether observed/test centre or randomly permuted groups) may have identical mean or median PRS values, but the proportion of individuals exceeding the threshold PRS may differ.

### Generalized linear model to predict the number of risk

A Generalized linear model (GLM) regression is used to predict the probability of developing CAD for every sample in the complete data set, as detailed in Section Study flow. When regressed on PRS and PCE, the model is:

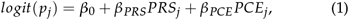

where *j ∈* (1, *J*) denotes the observation from the complete data set, *β*_0_ is the intercept, *β*_*PRS*_ and *β*_*PCE*_ are regression coefficients for the respective variables, and *p*_*j*_ is the predicted probability of developing CAD for the *j*_*th*_ individual.

The predicted incidence of CAD at the assessment level was calculated as the average of the predicted rates for all participants from the same assessment centre. As well as in models with PRS and PCE only, GLMs with BMI and TDI are also tested. Section Results for UKB assessment centres shows that the simple GLM has a very poor fit at the assessment centre level even though the individual level prediction is acceptable (based on the AUC results). The group level prediction became even worse after a new variable was added into the model. A better model is therefore needed to predict the group incidence rates and we introduce one in Section The Mundlak model to predict the number at risk.

#### Exploration of performance using simulated groups

To identify the reasons for the poor group-level predictions, we examined the performance of the same model on randomly labelled groups and specially designed groups. For the random casecontrol swaps groups, we assume that the complete data set has 9 groups of similar size, and then randomly label all samples in this complete data set from 1 to 9. The designed labelling method then randomly swaps cases and controls between groups to increase incidence heterogeneity. Table 2 lists the steps for the designed labelling method.

**Table 2.**
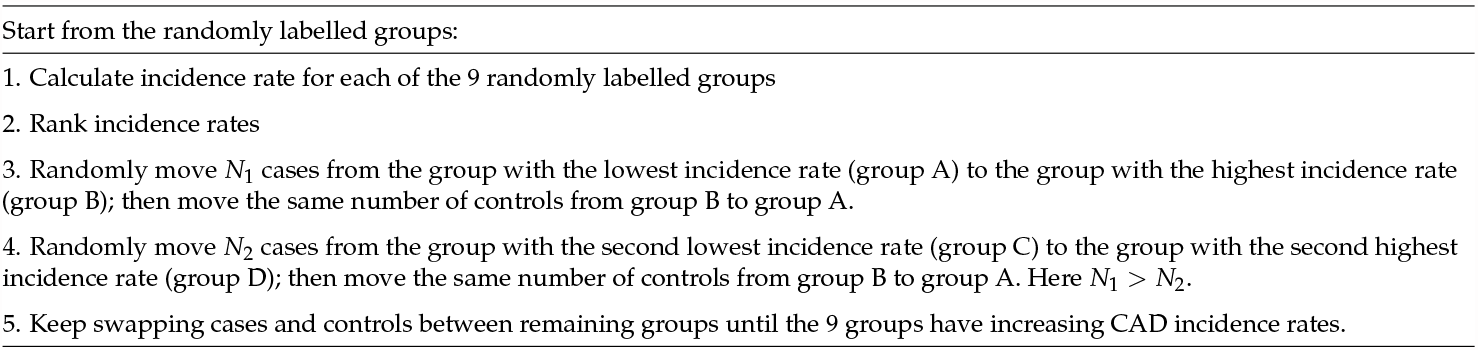
Random case-control swaps groups.

The results for both methods of simulated groups are reported in Section Results for simulated groups based on random case-control swaps. This result demonstrates the ability of GLM to distinguish between low and high incidence groups for the data set, which was created under the designed labelling method. Accordingly, we speculated that some cryptic group structure might play a role in the poor fitting at the assessment centre level, so we employed Pearson’s correlation tests to reveal the group structure. It turns out that the cryptic group structure in our data set is the reversed direction of the relationship of variables at the group level and the subgroup level. Such a scenario is common in statistical analysis and is referred to as Simpson’s paradox. Section Ascertainment bias confounds group rate estimation shows the detailed scenario in our data set.

### The Mundlak model to predict the number at risk

We are interested in developing a model to predict CAD risk at the assessment centre level without using the assessment centre label, so that this model can be used to estimate the number of CAD risks for a new group where we only observe covariates but not the group rate. Simple GLMs fit poorly at the assessment centre level, and this poor fit is due to the opposite directional effect of the variables at the group level and the subgroup levels (see results in Ascertainment bias confounds group rate estimation). A latent variable model cannot be used to predict the group rate because it requires an estimate of the group rate, such as the observed rate in a sample, from which the group-specific intercept term can be estimated.

The Mundlak model fits our needs well. This model was originally conceived by Mundlak in 1978 (Mundlak 1978) to analyse data consisting of repeated observations on economic units. In his model, group means of independent variables are included in addition to the original observed variables, so the assumption that observed variables should not be uncorrelated with unobserved variables is relaxed. Dieleman and Templin (2014) compared the random- and fixed-effects estimators (RE and FE, respectively) with the Mundlak model (called the within-between approach in this paper) for clustered data when unaccounted-for group-level characteristics affect the outcome variable. Even though RE and FE are commonly used competing methods in health studies, the Mundlak model outperforms those two estimators in their simulation study.

In this study, according to the GLM illustrated in Generalized linear model to predict the number of risk for regression on PRS and PCE, the Mundlak model simply adds group-mean variables into that model. We used the same approach as (Dieleman and Templin 2014):

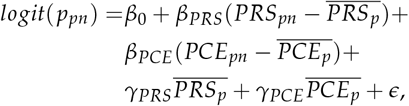

where *p ∈* (1…*P*) and *n ∈* (1…*N*) denote the group and observation identification within each group respectively and *P * N* = *J* from Equation 1. For the *n*_*th*_ individual belonging to the *p*_*th*_ assessment centre, 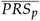 and 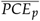 are the means of PRS and PCE for the *p*_*th*_ assessment centre. *β*_0_ is the intercept, *β*_*PRS*_ and *β*_*PCE*_ are regression coefficients for the group demeaned PRS and PCE, respectively, *γ*_*PRS*_ and *γ*_*PCE*_ are estimators for the corresponding group mean PRS and PCE, and *ϵ* is the residual. Here, Dieleman and Templin (2014) used the original variable minus the group-mean as the input variables, rather than the original variables, for reasons explained in Bell and Jones (2015). According to Dieleman and Templin (2014), every *β* represents the within-group effect and assesses changes within a group and every *γ* measures the effect of the corresponding variable between groups.

To quantify the uncertainty in the estimated incidence of CAD at the assessment centre level, we used the bootstrap method to establish a prediction interval. The bootstrap uses resampling techniques to create a list of test statistics of interest. The steps used are:

1. save the regression coefficients of the Mundlak GLM trained with the complete data set;
2. sample the same size of individuals with replacement from the complete data set;
3. calculate the new group mean of variables for this resampled data set from step 2;
4. apply the regression coefficients from step 1 to the step 2 data set and use the results to calculate the incidence of CAD for each assessment centre and save the results;
5. repeat steps 2-4 1000 times to get a list of estimated incidences of CAD at the assessment level.

The prediction 95% confidence intervals for each assessment centre then can be calculated.

### How the Mundlak model works

Results in Section The Mundlak model results show that the Mundlak model works well on prediction of CAD risk at the assessment centre level. The reason for this significantly improved performance is that group mean variables in the Mundlak model act as a proxy for unseen group specific behaviour, so the group structure can be captured in the Mundlak model. We next demonstrate a simulation experiment to better understand why the Mundlak model works.

The theory to support the simulation is that the risk of CAD increases with the increasing of PRS and PCE scores. We start the simulation with a reproduction of the Simpson’s paradox scenario, using the same complete data set as detailed in Section Study flow. Then we manually create groups based on which quantile one hidden variable falls in. We assume that this hidden variable *y*_*j*_ *∼ N*(*μ*_*j*_, 1) is a random variable with mean value calculated as a linear combination of *PRS*_*j*_ and *PCE*_*j*_, so that:

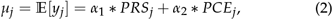

where *α*_1_ and *α*_2_ are correlation coefficients for variables PRS and PCE.

The correlation coefficients, *α*_1_ and *α*_2_, are used to determine the existence and extent of Simpson’s paradox. The severity of the reversed direction of the relationship of variables at the individual level and the group level can be controlled by the size of *α*_1_ and *α*_2_. For example, when *α*_1_ and *α*_2_ have the same signs, *Y* increases with increasing PRS and/or PCE. If we create several groups of equal size based on the ranked values of *y*_*i*_ from lowest to highest, so that the first group contains samples with the lowest values of *Y* and the last group contains samples with the highest values, then the first group should have the lowest average PRS *and* lowest average PCE and the last group the highest average PRS *and* highest average PCE. When GLM is regressed on PRS and/or PCE, individuals with higher values of PRS and PCE should have higher probability of developing CAD. Similarly, comparing groups with increasing values of PRS and PCE, the group with high PCE and PRS values has more risky individuals than the group with low values. In this case, the individual level and the group level have the same CAD rate trend, so Simpson’s paradox does not exist. However, predicted group rates will be biased towards the mean.

When the correlation coefficients have opposite signs, the relationship among groups is not as straightforward. For example, if we set *α*_1_ = − 0.5 and *α*_2_ = 1, *Y* decreases with increasing PRS, but increases with increasing PCE scores. We also create equal-sized groups based on the ranked values of *y*_*i*_ from lowest to highest. Under this scenario, the first group has the highest mean of PRS *and* the lowest mean of PCE, but the last group has the lowest mean of PRS *and* the highest mean of PCE. Because PRS and PCE contribute in opposite directions to group assignment, the CAD rates between groups will be less different than in the above scenario.

For the second scenario, because the risk of developing CAD depends on both PRS and PCE score in the same direction, GLM regressed on PRS and PCE will experience Simpson’s paradox, which will lead to poor predictive performance at the group level. But the Mundlak model accounts for the opposite direction by finding individual level and group level coefficients of opposite signs. Therefore, we expect a good fit of the GLM with the inclusion of group mean variables. Section Simulation results confirms this expectation.

## Results

### CAD events and rates

We extracted and compared the age of onset of the first CAD events between self-reported health outcomes with hospital inpatient data to determine the prevalence and incidence of CAD events. Supplementary Table 3 gives the number of first-ever CAD prevalence and incidence events from inpatient and selfreported records separately. Many participants reported CAD events in their self-reporting, but those events occurred too early to be recorded by inpatient data. This was consistent with suggestions from Eastwood *et al*. (2016) and Yeung *et al*. (2022), which both noted that using only UKB hospital inpatient data to identify prevalent cases would miss out many cases, as most prevalent cases were self-reported during the baseline assessment visit. Additionally, we found that the majority of participants with self-reported CAD events would have new CAD events recorded in their hospital inpatient records, with the majority occurring after they joined the UKB. Therefore, using only inpatient data would mistake actual prevalent cases as incidence cases. A total of 12 participants had only CAD events in their self-reported data and none in their hospital inpatient records, but no date of onset of CAD was given. We considered these participants as prevalent CAD cases. Thus, in conclusion, out of a total of 502,410 UKB participants, 16,558 participants had their first CAD event before they joined the UKB and 19,047 participants had their first CAD event after they joined the UKB.

**Table 3.** Summary statistics of risk factors for the complete data set. Risk factors in bold are parameters included in the calculation of PCE risk. HDL denotes high-density lipoprotein cholesterol.

| Risk factors | Overall (N = 263,087) | Males (N = 115,150) | Females (N = 147,937) |
| --- | --- | --- | --- |
| Incidence CAD events | 8,458 (3.21%) | 5,870 (2.22%) | 2,588 (0.99%) |
| **Age joined UKB** | 55.9 (39, 73) | 55.8 (39, 73) | 55.9 (40,70) |
| **Total cholesterol, mg/dl** | 226.7 (130, 320) | 222.5 (130,320) | 230.0 (130,320) |
| **HDL cholesterol, mg/dl** | 56.8 (20.3,100) | 50.4 (20.3,99.9) | 61.8 (20.3,100) |
| **Systolic blood pressure, mm Hg** | 137.2 (90,200) | 140.7 (90,200) | 134.5 (90,200) |
| **Smoking status %** | 43.0% | 47.6% | 39.4% |
| **Hypertension medication %** | 13.4% | 13.9% | 13.1% |
| **Diabetes mellitus %** | 1.3% | 1.8% | 1.0% |
| Body mass index | 27.0 (12.1, 66.2) | 27.4 (12.8, 61,7) | 26.7 (12.12, 74,7) |
| Townsend deprivation index | Calculated immediately prior to participant joining UKB.<br>Based on the preceding national census output areas. |  |  |

For all 22 assessment centres, we calculated age-standardized CAD prevalence rates on 1st October 2010 (the last day of attending assessment centre for all UKB participants) and non-standardized CAD incidence rates from 1st October 2010 to 30th September 2021 (the latest hospital inpatient record for CAD from our UKB file). Figure 2 (a) is the map for UKB assessment centres downloaded from UKB website and (b) and (c) are maps with CAD rates created following steps explained in Appendix UKB location co-ordinates. To calculate the age-standardized CAD prevalence in 2010, two additional steps were taken in addition to following the definition of CAD definition to distinguish between prevalence and incidence of CAD events. Firstly, we removed UKB participants who died after enrolment in the UKB but before 1st October 2010, and secondly, we redefined incidence CAD events that occurred after participants enrolled in the UKB but before 1st October 2010 as prevalence events.

**Figure 2.**
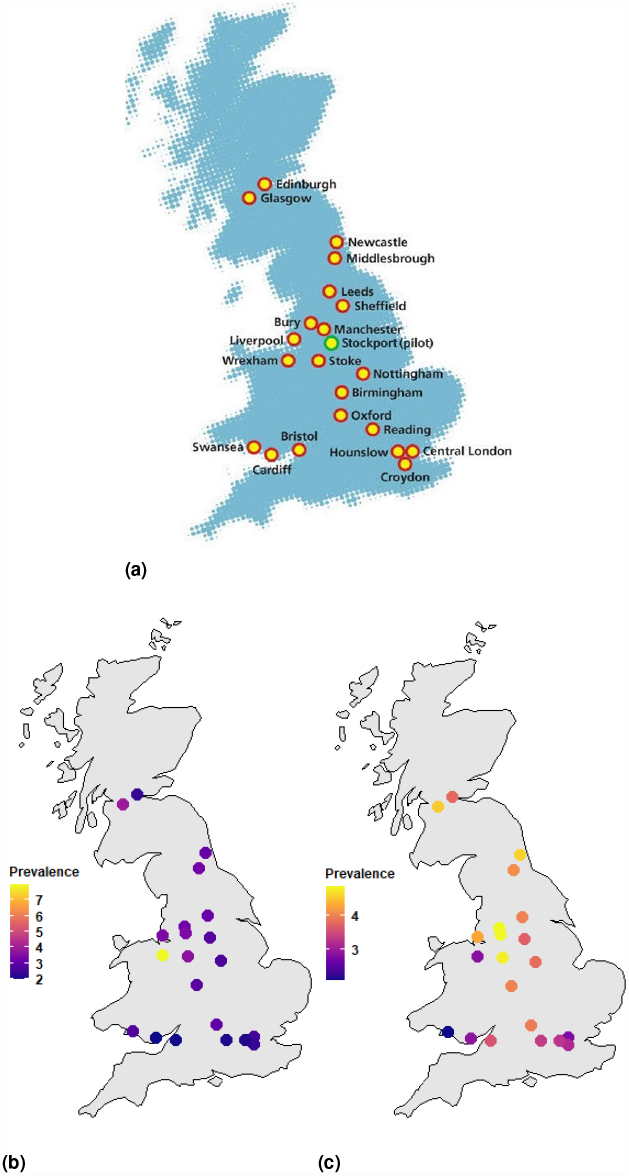
Maps and CAD rates of 22 UKB assessment centres (a) Locations of UK Biobank baseline assessment centres, (b) Age-standardized CAD prevalence rates, 2010, and (c) CAD incidence rates, 2010-2021.

Figure 2 (b) shows differences in the prevalence of CAD among centres. Cardiff and Bristol have the lowest CAD prevalence rates, whilst Wrexham and Glasgow have the highest rates. Figure 2 (c) shows CAD incidence (without age standardization) for each assessment centre. Stockport has the highest incidence rate, followed by Bury and Manchester. Among all 22 centres, Wrexham and Swansea were mobile assessment centres and Stockport was a pilot centre.

### Complete data set

After distinguishing prevalence and incidence CAD events, calculating PCE scores for eligible UKB participants, extracting the CAD-PRS, BMI and TDI provided by UKB, and filtering for samples with missing data, the complete data set had 263,087 participants, all of whom were White British. The detailed study flow chart is found in Section Study flow. The overall dataset had 3.21% incidence CAD event rate, with twice as many male patients as female patients. Summary statistics for risk factors used for PCE score calculation for men and women in the complete data set are found in Table 3, which lists the summary statistics (mean, minimum and maximum) for numerical risk factors and percentages for binary risk factors. In general, female participants had higher cholesterol levels, but lower levels of systolic blood pressure and BMI, and lower rates of smoking, hypertension medication and diabetes.

Figure 3 shows the density plots for PRS and PCE risk from the complete data set by CAD status and sex. Those plots show the ability of PRS and PCE risk to distinguish CAD cases and controls. The PRS density plots do not appear to differ between males and females, but samples with CAD from the complete data set have higher mean PRS values than samples without CAD.

**Figure 3.**
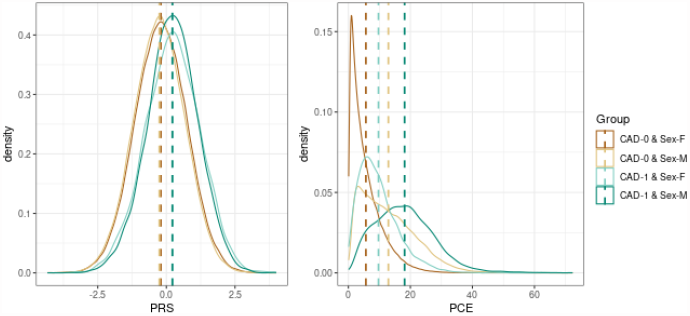
Density plots for PRS and PCE risk from the complete data set. CAD-0 denotes samples without incidence CAD events; CAD-1 denotes samples with incidence CAD events; F denotes female; M denotes male.

### Statistical tests results

The permutation test was used to compare the PRS distribution between groups because it makes no assumptions about the distributions and can capture differences in the tails of the PRS distributions. We first applied this test on PRS values between samples from England and Scotland, but didn’t find any significant differences. We then applied this test across UKB assessment centres, and plotted p-value results in a heat map. Figure 4 classifies the p-values of permutation tests between any two assessment centres into 3 groups. It is not a symmetrical heat map due to sampling error, as a permutation test is a Monte Carlo resampling test. The smaller the p-value in Figure 4, the higher the probability of a significant difference in PRS distribution between the two centres. For example, the distribution of PRS in Barts and Hounslow is different from many other assessment centres, but the distribution of PRS in Wrexham is not different from other locations.

**Figure 4.**
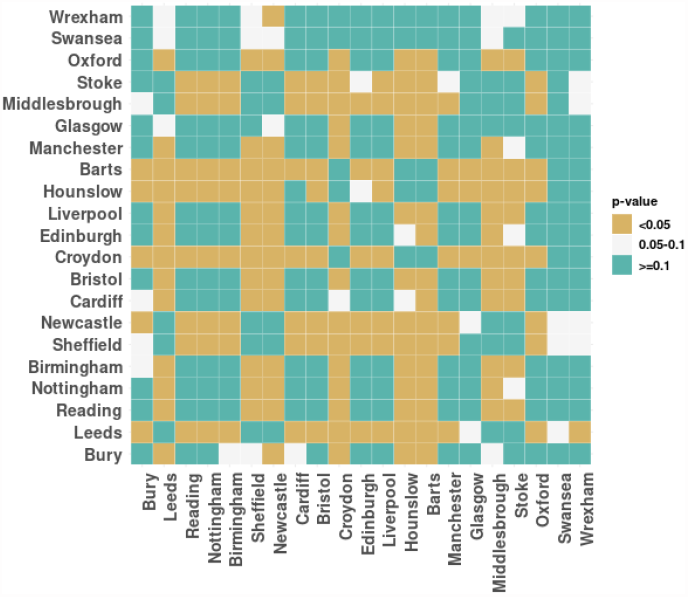
Permutation tests on PRS across UKB assessment centres. “*<* 0.05” denotes p-value less than 0.05; “0.05-0.1” denotes p-value between 0.05 and 0.1; “*≥* 0.1” denotes p-value greater than or equal to 0.1.

Figure 5 shows the p-values from permutation tests between any two assessment centres on the predicted disease risk obtained by the LTM based on PRS alone. The LTM estimates the proportion of individuals with a PRS greater than the LTM threshold PRS values from the entire UKB, which is a good proxy for predicted case rates, as two centres may have different proportions even if the mean PRS values between two centres are the same. The results in Figure 5 are similar to those in Figure 4, except for Wrexham and Swansea, where there is almost no difference in predicted disease risk between these two centres and the other centres. This is likely due to the small sample sizes.

**Figure 5.**
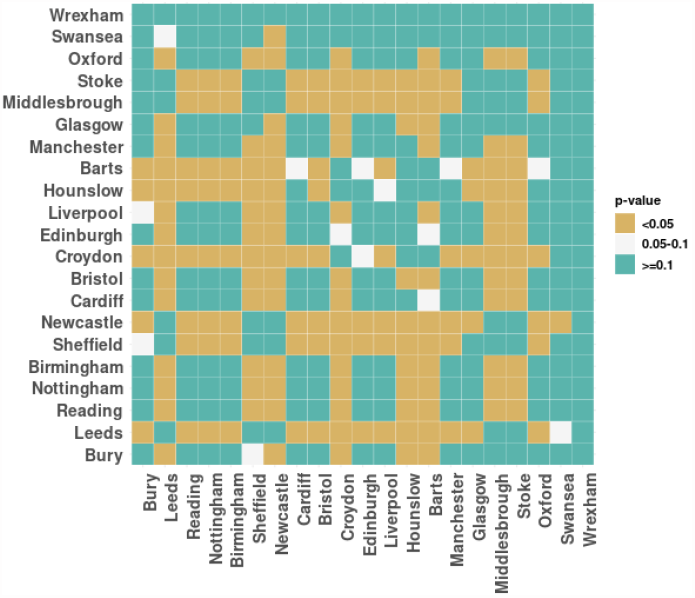
Permutation tests on the LTM predicted disease risk across UKB assessment centres. “*<* 0.05” denotes p-value less than 0.05; “0.05-0.1” denotes p-value between 0.05 and 0.1; “*≥* 0.1” denotes p-value greater than or equal to 0.1.

From the complete data set, we also compared the distribution of PCE risk between the England and Scotland samples using the Mann-Whitney test (Yang *et al*. 2021). We found significant distribution differences of PCE risk (p-value = 3.985e-05), rather than the small statistically significant differences found by (Yang *et al*. 2021) (p-value = 0.009) on the distribution of FRS and QRISK3 between England and Scotland. We then used the permutation tests on PCE risk across the UKB assessment centres, and the results are shown in Figure 6. Figure 6 shows that, with the exception of Wrexham, the PCE risk distributions of all the other centres are very different from each other.

**Figure 6.**
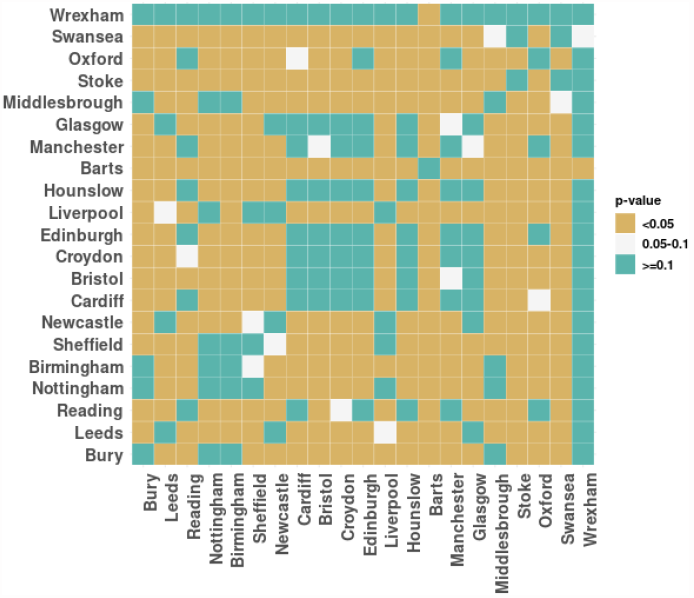
Permutation tests on PCE risk across UKB assessment centres. “*<* 0.05” denotes p-value less than 0.05; “0.05-0.1” denotes p-value between 0.05 and 0.1; “*≥* 0.1” denotes p-value greater than or equal to 0.1.

### Results from simple GLMs

#### Results for UKB assessment centres

We used GLMs regressed on selected variables to predicted the probability of developing CAD for each sample in the complete data set, and then calculated the assessment-level incidence of CAD as the mean of the predicted rates for all participants in the same assessment centre. Figure 7 plots the relationship between the observed case rates and the predicted cases rate from five GLMs and Table 4 gives the corresponding AUC from that GLM and the correlation between observed and predicted group rates. In general, the AUC is relatively high, especially when the PCE score is used independently. When we started with PRS and gradually added more variables in the model, the AUC increased slowly and all variables exhibited a significant positive relationship with the risk of CAD. However, the prediction of the case rate at assessment centre level is very poor, as the predicted case rates for all assessment centres are very close in all GLMs. The relatively high correlation of the PRS GLM is due to the fact that the predicted rate increases with the observed case rate, but the PRS line in Figure 7 shows that the predicted case rates remain very close across centres. We also tested GLMs with interaction and quadratic terms, but did not obtain better performance than for GLMs with only linear variables.

**Table 4.** Area under the curve (AUC) for GLMs regressed on the listed variables, trained and tested on the same complete data set, and the correlation between observed and predicted group rates. PCE denotes pooled cohort equation; PRS denotes polygenic risk score; BMI denotes body mass index; TDI denotes Townsend deprivation index

| Variables in GLM | AUC | Correlation |
| --- | --- | --- |
| PCE | 0.7303 | 0.2580 |
| PRS | 0.6318 | 0.6898 |
| PRS+PCE | 0.7515 | 0.3986 |
| PRS+PCE+BMI | 0.7523 | 0.3350 |
| PRS+PCE+TDI | 0.7523 | 0.2828 |
| PRS+PCE+BMI+TDI | 0.7532 | 0.2456 |

**Figure 7.**
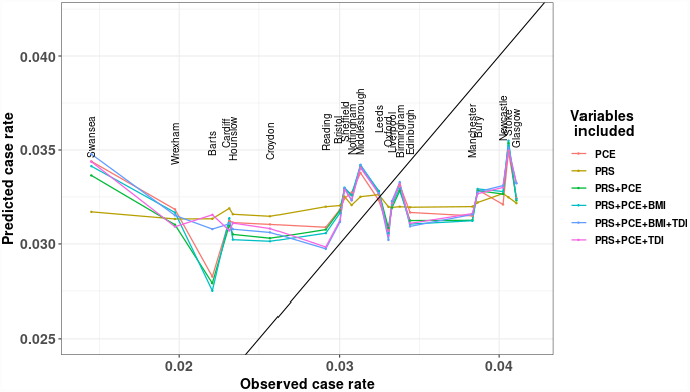
Predicted case rates from GLMs regressed on selected variables trained and predicted on the same complete data set. Observed case rates are CAD incidence rates for each UKB assessment centre and predicated case rates are the mean of the predicted rates for all participants in the same centre. PCE denotes pooled cohort equation; PRS denotes polygenic risk score; BMI denotes body mass index; TDI denotes Townsend deprivation index.

#### Results for simulated groups based on random case-control swaps

To identify the reasons for the poor assessment centre-level predictions, we manually created several groups with increasing case/control ratios using the method described in Section Exploration of performance using simulated groups. Figure 8(a) shows the results of predicted cases rates for 15 manually created groups, and 8(b) is modified from Figure 7 to have the same y-axis range as 8(a). Figure 8(a) shows a more obvious positive correlation between observed and predicted rates than 8(b), although the group prediction is still less satisfactory as the y-axis range is small. Based on this improved group-level prediction performance in Figure 8(a), we can infer that there exists some cryptic group structure that has played a role in the poor fitting at the assessment centre level in Figure 7.

**Figure 8.**
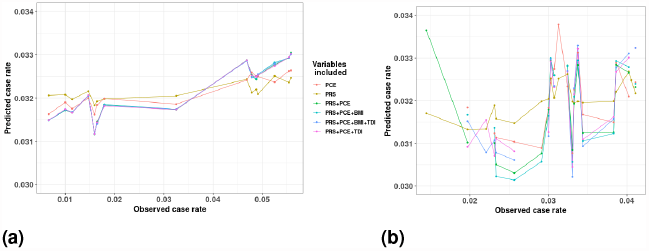
Predicted case rates on different groups from GLMs regressed on selected variables trained and predicted on the same complete data set for (a) manually created groups via random case-control swaps and (b) UKB assessment centres, modified from Figure 7 to have the same range of the y-axis as in (a).

#### Ascertainment bias confounds group rate estimation

We compared the correlation between two risk factors of the complete data set and at the group level using Pearson’s correlation. First, we calculated the correlation between each pair of variables for the complete data set following the flow chart detailed in Section Study flow and plotted the coefficients in a heat map (Figure 9(a)). For example, the coefficient between PRS and PCE risk was calculated using values from all 263,087 samples in the complete data set as 0.009. We then calculated the correlation between each pair of variables using the mean of the assessment centre for each variable. There are 21 assessment centres in the complete data set - we calculated the mean of each assessment centre for each variable. Using the same example, we had 21 PRS means and 21 PCE risk means, we then calculated Pearson’s correlation using these two sets of means. In this case, the correlation between PRS and PCE risk was 0.39, much higher than the previous value.

**Figure 9.**
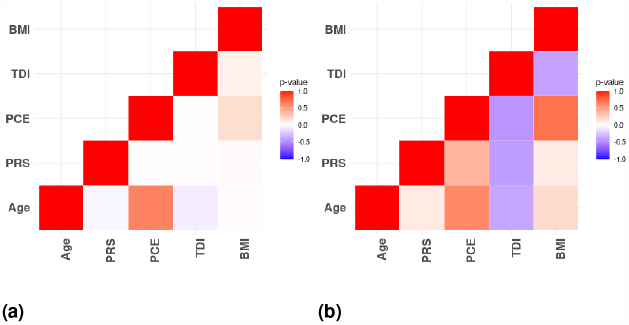
P-values from Pearson’s correlation tests (a) correlation for the complete data set and (b) Correlation using mean values from UKB assessment centres.

The inverse relationship between two variables at different levels is a well-known phenomenon, termed Simpson’s paradox (Pearl 2014). Another example in this study is the relationship between the variables PRS and TDI. We can see that their correlation is negative at the assessment centre level, but slightly positive for the complete data set. This reversed relationship can also be found for continuous variables included in the PCE risk calculation. For example, Figure 10 shows that such a phenomenon exists between PRS and HDL cholesterol levels and between systolic blood pressure and TDI. One possible reason for the inverse relationship at the individual level and group level is participation bias (Schoeler *et al*. 2023). For example, if a group has a higher rate of death from CAD for some reason (e.g. higher average age), then the group mean of the surviving people from whom a sample can be taken will have a lower PRS based risk, despite the homogeneity of genetics between the groups before people died off. This creates an ascertainment bias that varies between groups.

**Figure 10.**
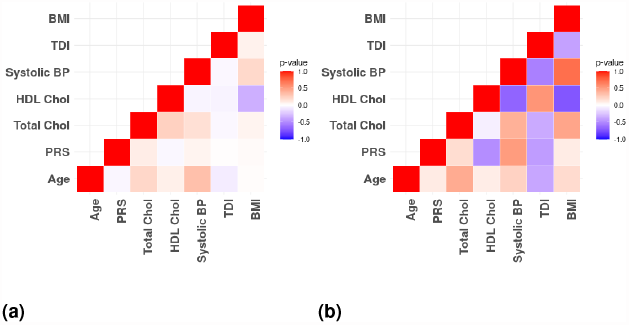
P-values from Pearson’s correlation tests (a) Correlation for the complete data set and (b) Correlation using mean values from UKB assessment centres.

### The Mundlak model results

After identifying the potential cause for the poor fit at the assessment centre level, we employed the Mundlak model to deal with this problem. The Mundlak model in this study was built based on GLMs regressed on the original variables as well as the group means of the same variables. Figure 11 shows a very strong positive relationship between the observed case rates and the predicted case rates for all 21 assessment centres after including the group mean variables in the GLMs. The AUCs from the Mundlak model are given in Table 5, where the values are very close to the AUCs in Table 4. The correlations between observed and predicted group rates from the Mundlak GLMs in Table 4 are higher than those in Table 4, except for the GLM regressed on PCE alone. This means that compared with simple GLMs, the Mundlak GLMs did not change the risk prediction at the individual-level, but significantly improved the prediction accuracy at the assessment centre level.

**Table 5.**
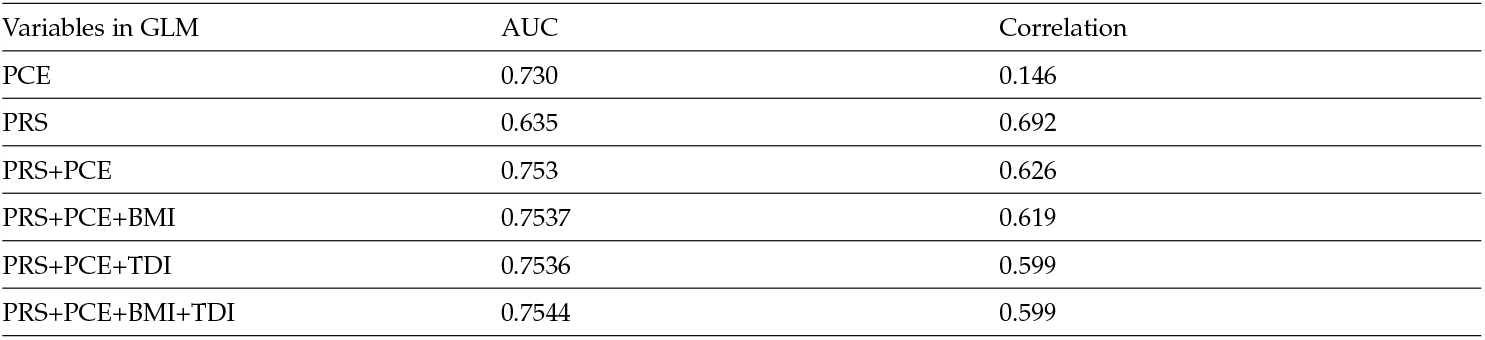
Area under the curve (AUC) for Mundlak models regressed on the listed variables, trained and tested on the same complete data set, and the correlation between observed and predicted group rates. PCE denotes pooled cohort equation; PRS denotes polygenic risk score; BMI denotes body mass index; TDI denotes Townsend deprivation index

**Figure 11.**
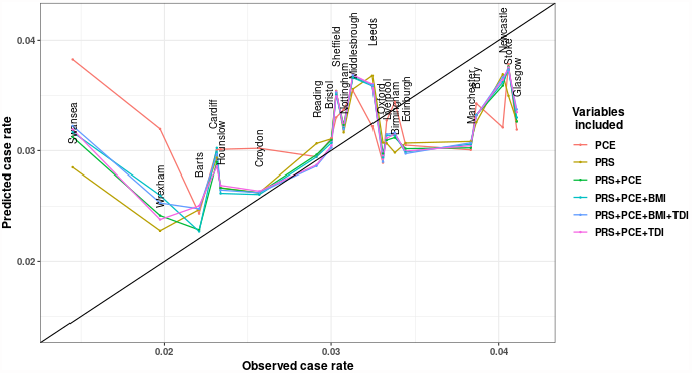
Predicted case rates from Mundlak GLMs regressed on selected variables trained and predicted on the same complete data set. Observed case rates are CAD incidence rates for each UKB assessment centre and predicted case rates are the mean of the predicted rates for all participants in the same centre. PCE denotes pooled cohort equation; PRS denotes polygenic risk score; BMI denotes body mass index; TDI denotes Townsend deprivation index.

The Swansea assessment centre is a notable outlier. A possible reason for this is that this centre has a larger number of older participants, as this centre has the highest average age of any centre in the complete data set (Supplementary Table 6). (Nanna et al. 2020) examined the performance of PCE in older adults, and found poor performance of PCE for ASCVD risk estimation in older adults. This centre has the highest mean value of PCE risk, but the lowest CAD incidence rate (Table 6 in the Supplementary material) and this phenomenon reduces the accuracy of the assessment centre level predictions. The relatively low mean PRS value in Swansea compared with other centres is consistent with its lower incidence of CAD, which also explains why the Mundlak model regressing on only PRS gives the closest predicted case rate to the observed case rate versus other models. Manchester and Glasgow are another two outliers and both centres have relatively higher CAD rates, but relatively low mean ages and PCE risk. One possible reason for the poor fit of Manchester and Glasgow can be explained by the high p-values from the permutation results shown in Figure 4 and Figure 5. The distribution of PRS in Manchester and Glasgow is not significantly different from PRS in other centres, which makes forecasting more difficult.

**Table 6.** Estimated Mundlak GLM coefficients along with standard errors, using de-meaned individual observations and groupmean variables.

|  |  |
| --- | --- |
| (Intercept) | −3.69*** (0.44) |
| PRS | 0.52*** (0.01) |
| PCE risk | 0.08*** (0.00) |
| Group Mean PRS | 3.90*** (0.57) |
| Group Mean PCE risk | 0.09 (0.04) |
| AIC | 68321.88 |
| BIC | 68374.28 |
| Log Likelihood | −34155.94 |
| Deviance | 68311.88 |
| Num. obs. | 263087 |

\*\*\* $p < 0.001$ ; \*\* $p < 0.01$ ; \* $p < 0.05$

The Mundlak GLM regressed on PRS and PCE risk has the highest correlation between the predicted case rates and observed case rates from Table 4. Table 6 lists the estimated coefficients for this model along with standard errors.

We used the bootstrap method to produce a 95% prediction interval for the Mundlak model regressed on PRS and PCE risk to quantify the uncertainty in the estimated incidence of CAD at the assessment centre level. Figure 12 shows that Wrexham has the widest confidence interval, followed by Swansea. Centres with predicted case rates close to observed case rates have relatively narrow prediction intervals.

**Figure 12.**
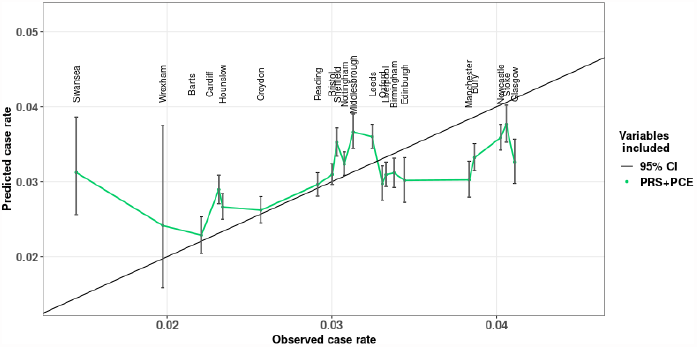
Predicted case rates and 95% prediction intervals from Mundlak GLMs regressed on PRS and PCE risk trained and predicted on the same complete data set. Observed case rates are CAD incidence rates for each UKB assessment centre and predicted case rates are the mean of the predicted rates for all participants in the same centre. PCE denotes pooled cohort equation; PRS denotes polygenic risk score.

Instead of using the PCE risk directly, we also tested the Mundlak GLM models on the PRS and the risk factors used in the PCE risk calculation. There are ten variables in total, including seven variables from PCE, PRS, BMI and TDI. We started by running Mundlak GLM on only one of the ten variables and selecting the model with the highest AUC, then we ran the Mundlak GLM on two variables (with 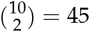 non-repeating combinations) and selected the two-variables Mundlak GLM with the highest AUC. The same steps were repeated, adding only one new variable each time, until all ten variables were included in the Mundlak GLM. Only linear combinations were included in the Mundlak GLM, because we checked that models with interactions or quadratic terms did not improve the predictive performance. We tested a total of 1023 Mundlak GLMs, and presented the results of the selected models in Figure 13 and Table 7. Among all ten variables, PRS has the best prediction power, followed by age and HDL cholesterol. Numbers in Table 7 are generally higher than in Table 5. To address any concerns around potential over-fitting due to the high number of regressors, we reassessed our models using leave-one-group-out cross-validation in Section Mundlak cross validation results.

**Table 7.** Area under the curve (AUC) for Mundlak GLMs regressed on the listed variables, trained and tested on the same complete data set, and the correlation between observed and predicted group rates. PRS denotes polygenic risk score; HDL denotes highdensity lipoprotein cholesterol; SBP denotes systolic blood pressure; TOT denotes total cholesterol; SMK denotes smoking status; HYS denotes hypertension status; DIA denotes diabetes status; BMI denotes body mass index; TDI denotes Townsend deprivation index

| Variables in Mundlak GLM | AUC | Correlation |
| --- | --- | --- |
| PRS | 0.635 | 0.692 |
| PRS+Age | 0.693 | 0.718 |
| PRS+Age+HDL | 0.732 | 0.688 |
| PRS+Age+HDL+SBP | 0.741 | 0.760 |
| PRS+Age+HDL+SBP+TOT | 0.748 | 0.767 |
| PRS+Age+HDL+SBP+TOT+SMK | 0.752 | 0.768 |
| PRS+Age+HDL+SBP+TOT+SMK+HPS | 0.755 | 0.852 |
| PRS+Age+HDL+SBP+TOT+SMK+HPS+DIA | 0.756 | 0.881 |
| PRS+Age+HDL+SBP+TOT+SMK+HPS+DIA+BMI | 0.757 | 0.882 |
| PRS+Age+HDL+SBP+TOT+SMK+HPS+DIA+BMI+TDI | 0.757 | 0.914 |

**Figure 13.**
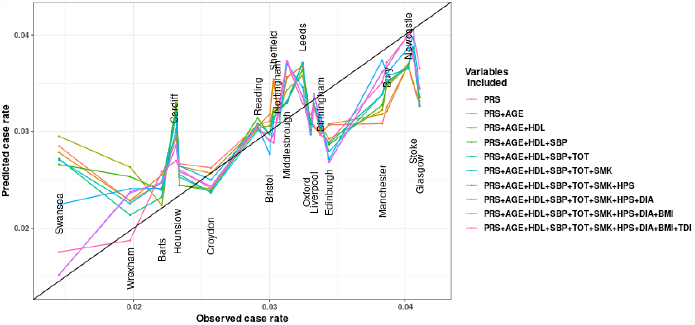
Predicted case rates from Mundlak GLMs regressed on PRS and other risk factors trained and predicted on the same complete data set. Observed case rates are CAD incidence rates for each UKB assessment centre and predicted case rates are the mean of the predicted rates for all participants in the same centre. PRS denotes polygenic risk score; HDL denotes high-density lipoprotein cholesterol; SBP denotes systolic blood pressure; TOT denotes total cholesterol; SMK denotes smoking status; HPS denotes hypertension status; DIA denotes diabetes status; BMI denotes body mass index; TDI denotes Townsend deprivation index.

#### Mundlak cross validation results

To assess out of sample performance, we applied the Mundlak model to the complete data set excluding one centre at a time, and then applied the model to the data set from this excluded centre. We called this method leave-one-centre-out cross-validation (LOCOCV) Mundlak GLMs. We averaged the predicted values for this one centre and obtained the predicted case rate for this centre. After applying the LO-COCV Mundlak GLMs to all centres, we thus obtained a list of predicted case rates for each centre, based on fitting the model to all other centres and the covariates.

Figure 14 and Figure 15 show the relationship between the observed rates and the predicted rates using PCE and the components of PCE respectively. Both figures show similar trends to Figure 11 and Figure 13, including the fact that Swansea is an obvious outlier, but Cardiff appears to be another more obvious outlier in Figure 14 and Barts is the largest outlier in Figure 15.

**Figure 14.**
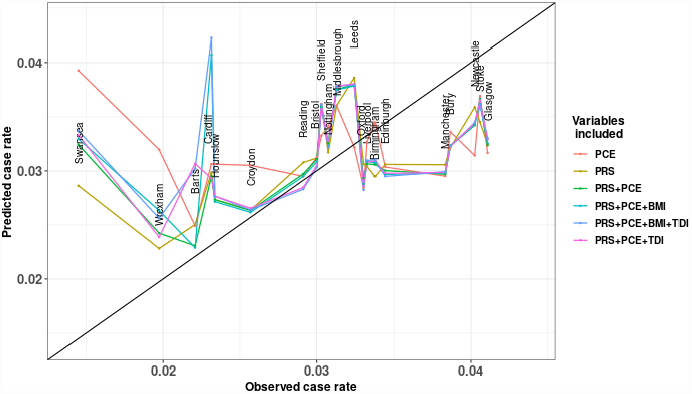
Predicted case rates from leave-one-centre-out crossvalidation (LOCOCV) Mundlak GLMs regressed on PRS, PCE and other factors. For each UKB assessment centre, the observed case rate is the CAD incidence rate, and the predicted case rate is that predicted by the Mundlak GLM trained on the data set excluding that centre. PCE denotes pooled cohort equation; PRS denotes polygenic risk score; BMI denotes body mass index; TDI denotes Townsend deprivation index.

**Figure 15.**
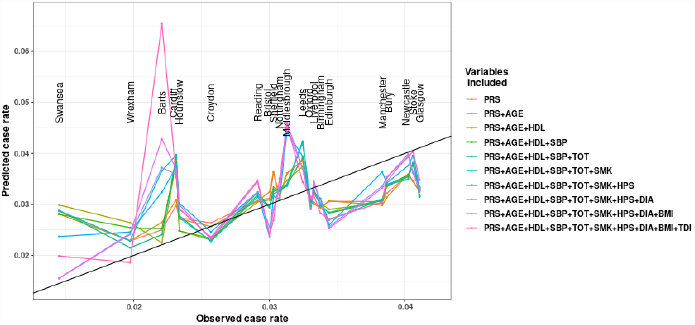
Predicted case rates from leave-one-centre-out crossvalidation (LOCOCV) Mundlak GLMs regressed on raw variables. For each UKB assessment centre, the observed case rate is the CAD incidence rate, and the predicted case rate is that predicted by the Mundlak GLM trained on the data set excluding that centre. PRS denotes polygenic risk score; HDL denotes high-density lipoprotein cholesterol; SBP denotes systolic blood pressure; TOT denotes total cholesterol; SMK denotes smoking status; HYS denotes hypertension status; DIA denotes diabetes status; BMI denotes body mass index; TDI denotes Townsend deprivation index.

Table 8 and Table 9 compare the correlation of observed and predicted group rates between a LOCOCV simple GLM and a LOCOCV Mundlak GLM regressed on PCE and variables included in PCE, respectively. In Table 8, the correlation from the Mundlak model is always higher than the corresponding correlation from the simple GLM, except for the model regressed on PCE risk only. In Table 9 such an exception happens when TDI is added to the model.

**Table 8.** The correlation between observed and predicted group rates from leave-one-centre-out cross-validation (LOCOCV) simple GLMs and LOCOCV Mundlak GLMs regressed on PRS, and other risk factors. PCE denotes pooled cohort equation; PRS denotes polygenic risk score; BMI denotes body mass index; TDI denotes Townsend deprivation index

| Variables | LOCOCV GLM | LOCOCV Mundlak GLM |
| --- | --- | --- |
| PCE | 0.138 | 0.014 |
| PRS | 0.251 | 0.619 |
| PRS+PCE | 0.304 | 0.498 |
| PRS+PCE+BMI | 0.245 | 0.274 |
| PRS+PCE+TDI | 0.133 | 0.409 |
| PRS+PCE+BMI+TDI | 0.109 | 0.158 |

**Table 9.** The correlation between observed and predicted group rates from leave-one-centre-out cross-validation (LOCOCV) Mundlak GLMs regressed on PRS, and other risk factors. PRS denotes polygenic risk score; HDL denotes high-density lipoprotein cholesterol; SBP denotes systolic blood pressure; TOT denotes total cholesterol; SMK denotes smoking status; HYS denotes hypertension status; DIA denotes diabetes status; BMI denotes body mass index; TDI denotes Townsend deprivation index

| Variables | LOCOCV GLM | LOCOCV Mundlak GLM |
| --- | --- | --- |
| PRS | 0.251 | 0.619 |
| PRS+Age | -0.053 | 0.585 |
| PRS+Age+HDL | 0.297 | 0.497 |
| PRS+Age+HDL+SBP | 0.289 | 0.531 |
| PRS+Age+HDL+SBP+TOT | 0.301 | 0.473 |
| PRS+Age+HDL+SBP+TOT+SMK | 0.316 | 0.310 |
| PRS+Age+HDL+SBP+TOT+SMK+HPS | 0.326 | 0.497 |
| PRS+Age+HDL+SBP+TOT+SMK+HPS+DIA | 0.290 | 0.531 |
| PRS+Age+HDL+SBP+TOT+SMK+HPS+DIA+BMI | 0.221 | 0.413 |
| PRS+Age+HDL+SBP+TOT+SMK+HPS+DIA+BMI+TDI | 0.252 | 0.186 |

### Simulation results

To better understand why the Mundlak model gave much better predictive performance at the assessment centre level, we designed a simulation experiment as described in Section How the Mundlak model works. Following the simulation design, we assumed that there was a hidden random variable with mean value calculated as a linear combination of PRS and PCE risk and then manually created 9 groups based on which quantile the hidden variable fell into.

If *α*_1_ and *α*_2_ from Equation 2 were assumed to have the same sign, the group with fewer CAD events should have lower values of PRS and PCE risk. We set *α*_1_ = *α*_2_ = 0.2 and then compared the performance of the simple GLMs when regressed on PRS only and and the performance of the Mundlak GLM when regressed on PRS and group mean PRS. The blue and green lines from Figure 16(a) show that the in-sample Mundlak model has much better group-level prediction than the in-sample GLM. The red line from Figure 16(a) shows that even the leave-one-group-out cross validation method has much better performance than the naive model. This is because, in the simulation setting, the observed case rate was determined by both PRS and PCE risk, so regressing on PRS alone could not predict the case rate well. When only PRS was included in the Mundlak model, the dependence of CAD on PCE could be captured by the group mean of PRS, so the Mundlak model should perform much better than a simple GLM. The group mean of the variable acts as a proxy for unseen group-specific behaviour in the Mundlak model.

**Figure 16.**
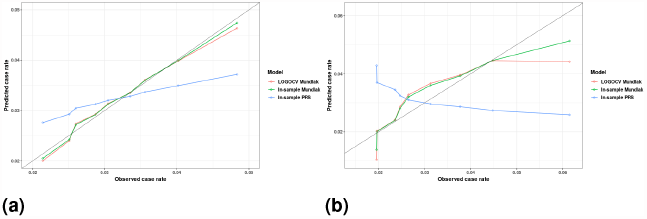
Predicted case rates for manually created groups, (a) when *α*_1_ and *α*_2_ from Equation 2 have same signs and (b) when *α*_1_ and *α*_2_ from Equation 2 have opposite signs. LO-GOCV denotes leave-one-group-out cross-validation.

If *α*_1_ and *α*_2_ from Equation 2 are assumed to have opposite signs and the groups were still determined by the hidden variable in Equation 2, there is no simple relationship between the severity of CAD risk and the value of PRS and PCE risk. We set *α*_1_ = *−*0.5 and *α*_2_ = 1 and let the GLM and Mundlak GLM both be regressed on PRS only. In this setting, the groups were determined by the opposite direction between PRS and PCE risk, but the risk of CAD was dependent on both variables in the same direction, so regressing CAD only on PRS experiences the Simpson’s paradox. Figure 16(b) shows the results from the in-sample GLM (blue line), the in-sample Mundlak GLM (green line) and the LOGOCV Mundlak GLM (red line). The in-sample GLM actually predicts low case rates for groups with high observed case rates and predicts high case rates for groups with low observed case rates. The Mundlak GLM can reveal the hidden inverse relationship between PRS and PCE risk, because group mean PRS acts as a proxy for the unseen relationship.

## Discussion

We proposed a framework for estimating the CAD case rate or number at risk in a homogeneous group of people, based on combining genetic and non-genetic contributions to risk. We demonstrated that simply fitting a logistic regression to the UK Biobank and then estimating group rates as the average predicted probability of CAD in the target sample has exceptionally poor performance. We showed that this is largely attributable to a reversal of correlation between genetic and non-genetic risk factors at the group or cohort level compared to the correlation at individual level. Such behaviour manifests as an example of Simpson’s Paradox wherein, for example, PRS and TDI are positively correlated across participants at the individual level, but the group specific mean values are negatively correlated. This can occur due to ascertainment bias, also known as participation bias or collider bias.

Population-based cohort studies, including the UKB study, are subject to participation bias. Fry *et al*. (2017) compared the sociodemographic and health-related characteristics of UKB participants with the general population and found that the UKB participants were more likely to be older, female and wealthier. Weng *et al*. (2019) compared the TDI gathered from 8,848 households in the 2001 UK Census and the 502,625 participants in the UKB cohort and found that UKB participants were generally less deprived than the general UK population. Schoeler *et al*. (2023) demonstrated that the selective participation of the UKB cohort twisted the genome-wide associations and genetic correlation results compared with results in probability samples. Our study showed that participation bias altered risk prediction at the group level. Using the same UKB data, the effect of population structure in different geographical areas has been studied by Lin *et al*. (2022) on the estimation of SNP heritability. Our study discussed the effect of PRS in different regions.

An example of a cause of such bias is where two groups of initially similar polygenic risk score distributions experience different CAD rates due to an unobserved or lurking variable such as a differing age profiles or lifestyle factors. Since PRS also contributes to risk, the higher death rate in one group will lead to the survivors having lower average polygenic risk scores as more of those with higher polygenic risk will have died and been removed as candidates for a sample in that group. Thus the samples from the two groups will have lower polygenic risk in the group with higher case rates. The direction of the relationship between polygenic risk and disease status is then reversed at the group (as opposed to individual) level.

We showed that this source of bias exists within the UK Biobank when using the assessment centres as groups, but then showed how to account for this structure using a Mundlak model wherein group specific means of covariates are included in the regression model. We demonstrated that such an approach has the ability to predict individual level disease status, with an accuracy that is improved relative to a model without such terms. But more importantly it has much improved ability to estimate the number at risk or case rate of a prospective group using samples for which disease status is unknown, but the regression covariates are available. UKB assessment centres have been used to adjust for bias in statistical analysis. For example, Lu *et al*. (2022) conducted GWAS and constructed PRS using UKB data, adjusting for terms such as age, sex and recruitment centre in their models.

Recent research (Lin *et al*. 2023) using the same UKB data suggests that factors such as age, sex, genetic batch, and assessment centre potentially exert a greater influence on PRS predictions compared to the inclusion of principal components (PCs); whereas including the top 10 (or more) PCs is the current approach used to adjust PRS predictions at the individual level in the presence of genetic heterogeneity. Our study shows that adding group specific means of covariates can also improve prediction at the individual level.

Most compelling of all is our result that the Mundlak regression model performs consistently well on out-of-sample group rate predictions as evidenced by the leave-one-group-out crossvalidation. This demonstrates that the ascertainment (or participation or collider) bias that causes the individual level logistic regression model to perform poorly in group-rate predictions is reduced in a systematic, consistent, and appropriate manner across assessment centres. This, in contrast to a latent variable or mixed model with group-specific intercept terms, can be used to predict group-rates based on new samples without an existing and accurate estimate of disease case rates. In our simulation experiment, we have shown that the Mundlak model can reveal the hidden inverse relationship between PRS and PCE risk even when only PRS was included in the model. This suggests that the Mundlak model has the potential to make accurate predictions when there is a significant variable that determines the risk but cannot be incorporated into the model directly.

Commercial genetic testing services have been sold more than 27 million times, but the ability of genetic factors to assess risk did not outperform common methods for CAD (van Dam *et al*. 2023). They also pointed out that risk assessment for CAD based on simple questionnaires or variables from electronic health records is as good or better than risk prediction based on genetics alone. For this reason, they recommended continuing to use questionnaire techniques for initial risk assessment rather than relying on genetic testing alone to determine risk. Our results suggest that for commercial providers of genetic testing services, prediction at the individual level can be significantly improved by adding group mean variables to the risk prediction model, and that age is a relatively easily obtained group indicator.

This study has limitations. The first limitation is that we use the group specific means of the same variables that are used in simple GLMs to adjust for the ascertainment bias. The group means of the independent variables may not fully capture the ascertainment bias between centres, as there may be other characteristics at the assessment centre level that affect the outcome, but that we haven’t included in our analysis. As health facilities in a single geographical area may share budgets, Dieleman and Templin (2014) noted that other sources may introduce ascertainment bias to health facilities, including guiding policies, attitudes towards treatment, population, disease patterns and supply constraints. For the UKB assessment centres, the original function of each assessment centre (for example, whether it is a clinic or a hospital), is another possible characteristic. If we had more centre-specific variables to add to this model, it might help explain more of the variation. Fortunately, if any such unseen factors are in any way correlated with any variables we do include at the group level, then the Mundlak model will account for them, up to that level and correlation.

Additionally, we only test the Mundlak model on the UKB participants, not on other external data sets. Single ancestry basis is another limitation of this study, as the complete data set only includes White British. Many studies have called for an increase in diversity in large-scale genetic association studies (e.g. Duncan *et al*. (2019), Schoeler *et al*. (2023)). Also, the accuracy of disease risk prediction was shown to improve after adding family history to the model (Gim et al. 2017), but this study did not explore the effect of family history on the group structure or the effect of other risk factors from the UKB resources. This can be investigated in future studies.

## Conclusions

We distinguished prevalence and incidence CAD events for all UK Biobank participants and identified geographical variations in CAD age-standardized rates across UKB assessment centres. The standard CAD-PRS provided by the UKB resources was selected to represent the genetic risk, as this set of PRS had the best predictive performance. We calculated PCE risk to represent the non-genetic risk factors for CAD. There were significant distributional differences in PRS and PCE risk between UKB participants from England and Scotland, according to the results of the Mann-Whitney test. Permutation test results showed that PRS from different assessment centres differed significantly. The group level predictive performance of simple GLMs was biased by a reversal of the correlation between genetic and non-genetic risk factors at the group or cohort levels, compared to the individual level. This behaviour was effectively modified by the Mundlak model, which included the group specific means of covariates along with the original covariates in GLMs. The group means of the covariates acted as a proxy for the unobserved group-level characteristics that affected the outcome variables. The Mundlak model has the advantage of predicting the number at risk in a new group, given a sample of individual-level data. We showed that our model can effectively predict case rates in out-of-sample groups even in the presence of ascertainment bias that confounds group rate estimation. Our method corrects for systematic biases at the cohort level and has potential applications in public health planning, including screening programmes and early intervention strategies.

## Appendix

### Results for age groups

Splitting groups by age is common in health-related studies, so we repeated our analysis using age as the group indicator. Figure 17 shows that PCE risk and PRS have a strong negative correlation for age groups. This is because participants with highly elevated PRS had developed CAD or other diseases, so they don’t show up in the complete data set, confirming the existence of collider bias. As there were only few participants in the age group [35,39], we excluded this group from the analysis.

**Figure 17.**
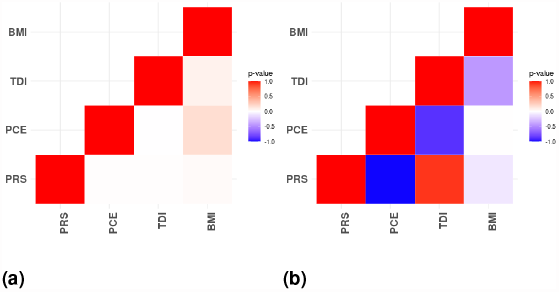
P-values from Pearson’s correlation tests: (a) correlation for the complete data set and (b) correlation using mean values from age groups.

Figure 18 and Figure 19 show the predicted case rates plotted against the observed case rates from the simple GLMs and the Mundlak GLMs respectively. When the PCE risk is included in the regression model, both the individual and group level perform well as age is included in the calculation of the PCE risk. Table 10 shows that the simple GLMs and the Mundlak GLMs have similar levels of AUC. The advantage of the Mundlak model is evident when regressing only on the PRS (Figure 19), as the predicted case rates are close to the observed case rates, but not in the simple GLM (Figure 18).

**Table 10.** The area under the curve (AUC) for each Mundlak model trained and tested on the same complete data set. PCE denotes pooled cohort equation; PRS denotes polygenic risk score; BMI denotes body mass index; TDI denotes Townsend deprivation index.

| Variables in GLM | AUC - simple GLMs | AUC - Mundlak GLMs |
| --- | --- | --- |
| PCE | 0.7303 | 0.7318 |
| PRS | 0.6318 | 0.6874 |
| PRS+PCE | 0.7515 | 0.7526 |
| PRS+PCE+BMI | 0.7523 | 0.7534 |
| PRS+PCE+TDI | 0.7523 | 0.7534 |
| PRS+PCE+BMI+TDI | 0.7532 | 0.7541 |

**Figure 18.**
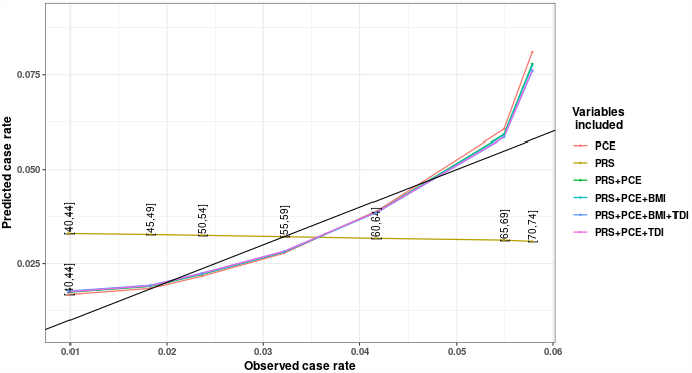
Predicted case rates from GLMs regressed on selected variables trained and predicted on the same complete data set. Observed case rates are CAD incidence rates for each age group and predicated case rates are the mean of the predicted rates for all participants in the same age group. PCE denotes pooled cohort equation; PRS denotes polygenic risk score; BMI denotes body mass index; TDI denotes Townsend deprivation index.

**Figure 19.**
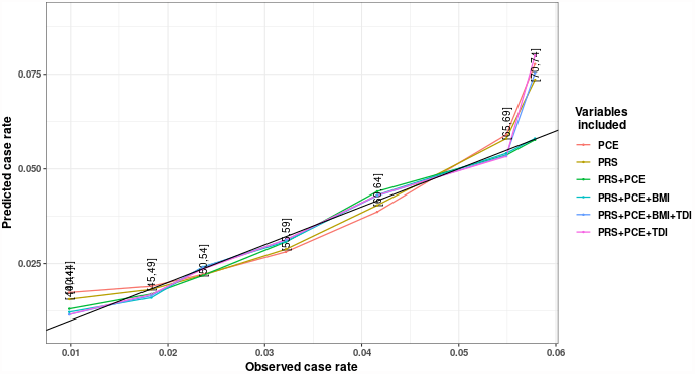
Predicted case rates from Mundlak GLMs regressed on selected variables trained and predicted on the same complete data set. Observed case rates are CAD incidence rates for each age group and predicted case rates are the mean of the predicted rates for all participants in the same group. PCE denotes pooled cohort equation; PRS denotes polygenic risk score; BMI denotes body mass index; TDI denotes Townsend deprivation index.

The prediction intervals in Figure 20 were generated using the method described in Section The Mundlak model to predict the number at risk with PRS as the only input variable. We trained both models on a subset of the complete data set, where the subset contained 70% of the randomly selected samples from each age group. We used the same age groups as in Table 11, but removed age group [35,39] as there were only 3 participants in this group. The test data set then contained 30% of the randomly selected samples from each age group. The trained models were applied to the test data set. This process was repeated 1000 times to obtain the confidence intervals.

**Table 11.** Age groups with proportions. ESP denotes European standard populations. Figures in the 2013 ESP % column are published proportions for each age group from the 2013 ESP distribution. Figures in the adjusted 2013 ESP % column are adjusted from the 2013 ESP % column so that the sum of the proportions in the study equals 1

| Age group | 2013 ESP % | Adjusted 2013 ESP % |
| --- | --- | --- |
| [35,39] | 0.070 | 0.137 |
| [40,44] | 0.070 | 0.137 |
| [45,49] | 0.070 | 0.137 |
| [50,54] | 0.070 | 0.137 |
| [55,59] | 0.065 | 0.127 |
| [60,64] | 0.060 | 0.118 |
| [65,69] | 0.055 | 0.108 |
| [70,74] | 0.050 | 0.099 |

**Figure 20.**
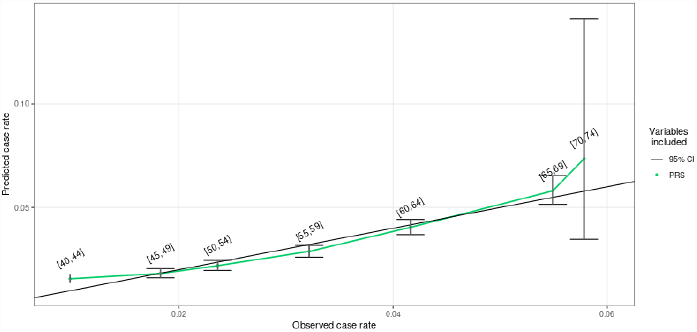
Predicted case rates and 95% prediction intervals from Mundlak GLMs regressed on PRS trained and predicted on the same complete data set. Observed case rates are CAD incidence rates for each age group of UKB participants and predicted case rates are the mean of the predicted rates for all participants in the same group. PRS denotes polygenic risk score.

Comparing the GLM and the Mundlak GLM regressed on PRS, the Mundlak GLM has a better risk classification performance, with an net reclassification improvement (NRI) of 3.54% (95% CI, 2.12% to 4.92%). This result is similar to the NRI obtained by Elliott *et al*. (2020) by comparing the model with PCE and PRS with the model with PRS only.

van Dam *et al*. (2023) showed that the incidence in the 10% most at risk group of individuals increased from 2.4-fold and 3-fold to 4.7-fold risk for CAD by including common risk factors in the model with PRS only. Our results showed that the incidence in the 10% most at risk group of individuals increased from 2.3 (95% CI, 2.1 to 2.5) to 2.9 (95% CI, 2.7 to 3.1) times the risk of CAD by including the group mean PRS in the model with PRS only.

### LDpreds CAD-PRS calculation

To calculate CAD-PRS, Privé *et al*. (2020a) restricts the UKB participants to unrelated and white-British in several steps. Privé *et al*. (2020a) first selects individuals whose genotype data are used to compute the principal components (PCs) in the UKB (Data field 22020). Detailed information on the quality control procedure for performing the PC analysis is described in section S3 of Bycroft *et al*. (2018). Secondly, they compute a robust Mahalanobis distance based on the first 16 PCs on the individuals selected in the first step, and further restrict individuals to those within a log-distance of 5, the threshold used by Privé *et al*. (2020b). After this step, a set of genetically homogeneous individuals is obtained. Finally, they restrict the SNPs to the HapMap3 variants used in PRS-CS (Ge *et al*. 2019). Privé *et al*. (2020a) obtains a cohort of 362,320 individuals and 1,117,493 variants. We repeat their process and obtain a slightly smaller sample size of 362,263 (withdrawal of some UKB participants) and exactly the same SNP size of 1,117,493.

LDpred2 obtains joint effects from externally published summary statistics and a correlation matrix, and then uses Gibbs sampling to obtain the posterior mean effect sizes. LDpred2 computes 4 sets of PRS using different parameter selection options. We only select the set with the highest prediction accuracy (SNP-based heritability is 11%) on the validation set (352,263 individuals) when 10,000 individuals are selected to train the model.

### UKB location co-ordinates

UKB provides the grid coordinates for all assessment centres (UKB Resource 11002). These grid coordinates are not latitude and longitude information, but figures obtained from the Ordnance Survey National Grid geographical reference system, whose measurements are easting and northing with a reference point near the Isles of Sicily (UK Biobank: deriving the grid coordinates). We first translated the UKB grid coordinates of the UKB into latitude and longitude information, and then used these to create CAD rate maps.

### Age standardized rates

UKB participants were enrolled between the ages of 37 and 73. To generate age-standardized CAD prevalence rates, we first converted the original proportions for 8 age groups from the 2013 European standard populations distributions into adjusted proportions to make the total proportion equal to one. The age-standardized prevalence for each assessment centre is calculated as the sum of the adjusted prevalence from each age group. The adjusted prevalence is the original prevalence for each age group multiplied by the corresponding adjusted 2013 ESP proportions. Table 11 shows the age groups and the corresponding proportions.

## Data availability

The study analyses were based on data from the UK Biobank website (http://www.ukbiobank.ac.uk). UK Biobank data is open source and available to researchers following acceptance of a research proposal and payment of an access fee.

## Funding

This publication has emanated from research conducted with funding from the Science Foundation Ireland under Grant number [SFI/12/RC/2289_P2]. For the purpose of Open Access, the author has applied a CC BY public copyright licence to any Author Accepted Manuscript version arising from this submission.

## Conflicts of interest

The authors declare no competing interests.

## Notes

### Competing Interest Statement

The authors have declared no competing interest.

### Author Declarations

The study uses data that has been published and used previously from http://www.ukbiobank.ac.uk

